# Salt intake and the risk of NAFLD and NASH: A prospective cohort and Mendelian randomization study

**DOI:** 10.1101/2025.05.27.25328391

**Authors:** Shun Li, Hao Wang, Xiao-Qian Xu, Wei-Ming Li, Hong You, Ji-Dong Jia, You-Wen He, Yuan-Yuan Kong

## Abstract

**Background:** While excess salt intake is known to affect cardiovascular health, its role in nonalcoholic fatty liver disease (NAFLD) and nonalcoholic steatohepatitis (NASH) is less established. We aimed to examine the longitudinal association between salt intake and incident NAFLD/NASH.

**Methods and findings:** This study included 494,170 UK Biobank participants without NAFLD/NASH at baseline. Salt exposure was assessed via self-reported salt-adding frequency (four-point Likert scale) and estimated 24-hour sodium intake from spot urine. Incident NAFLD/NASH cases were defined by diagnostic codes. Hazard ratios (HRs) and 95%CIs were calculated by using Cox proportional hazards models. Mendelian randomization and mediation analyses were conducted to infer causality and explore underlying mechanisms. Over a mean 14.3-year follow-up, 7,307 NAFLD and 630 NASH cases were identified. Both higher salt-adding frequency and sodium intake were significantly associated with elevated NAFLD/NASH risk. Compared to those who never/rarely added salt, adjusted HRs (aHRs) for NAFLD were 1.02 (95% CI: 0.98–1.10), 1.20 (1.09–1.31), and 1.31 (1.16–1.48) for sometimes, usually, and always, respectively (*P*-trend < 0.001). Always adding salt was also linked to higher NASH risk (aHR = 1.42; 95% CI: 1.01–1.99). Per 1g increase in estimated 24-hour sodium intake, NAFLD and NASH risks increased (aHRs = 1.82; 95% CI: 1.73–1.92 and 2.17; 95% CI: 1.81–2.61). Genetically predicted salt-adding frequency was also associated with increased NAFLD risk (Odds ratio = 1.54; 95% CI: 1.16–2.05). The salt-NAFLD associations were more pronounced among individuals with normal body-mass index (BMI) or normal alanine transaminase (*P*-interaction = 0.002, 0.005), with BMI mediating 16.4% (95% CI: 13.1–22.4%) of the salt–NAFLD association.

**Conclusions:** Higher salt intake is independently associated with increased NAFLD/NASH risk, particularly in metabolically healthy individuals. These findings support liver-focused strategies in salt reduction policies.

## Introduction

Non-alcoholic fatty liver disease (NAFLD) is the most prevalent chronic liver condition globally, affecting approximately 30% of the population, including a rising number of younger individuals [1]. NAFLD encompasses a spectrum of liver abnormalities, from simple steatosis to non-alcoholic steatohepatitis (NASH), which can progress to cirrhosis, hepatocellular carcinoma, and liver-related mortality [2]. In the absence of approved pharmacological treatments, lifestyle modification remains the cornerstone of NAFLD management [3]. However, identifying modifiable dietary contributors to NAFLD, particularly in the general population, remains a key challenge.

Excess dietary sodium has long been implicated in the pathogenesis of hypertension and other cardiovascular diseases [4], yet emerging evidence suggests that its detrimental effects may extend beyond cardiovascular health, including associations with all-cause mortality [5]. Animal studies indicate that high sodium consumption can induce hepatic steatosis [6], and high sodium intake may drive some NAFLD risk factors such as obesity [7]. A recent meta-analysis, mostly based on cross-sectional data, reported a 60% higher NAFLD risk in individuals with high versus low sodium intake, while also emphasizing the need for well-designed studies to validate these findings [8]. Nonetheless, evidence from large-scale prospective studies remains scarce, and the potential causal relationship between salt intake and NAFLD has not been fully established. Furthermore, it remains unclear whether this association is more evident for certain population subgroups or the pathway of it.

To address these gaps, we investigated the association between salt intake—captured through both self-reported frequency of adding salt to foods and biomarker-based estimates of 24-hour sodium intake—and the incidence of NAFLD and NASH in the UK Biobank, a large prospective cohort. To strengthen causal inference, we performed Mendelian randomization (MR) analyses using genetic instruments for salt-adding frequency. We also explored the potential mediating role of body mass index (BMI) using both observational and MR-based mediation approaches. Additionally, subgroup and sensitivity analyses were conducted to evaluate effect modification and test the robustness of our findings (Figure 1). This study aims to clarify the role of dietary salt in the development of fatty liver disease and inform evidence-based strategies for NAFLD prevention.

**Fig 1.**
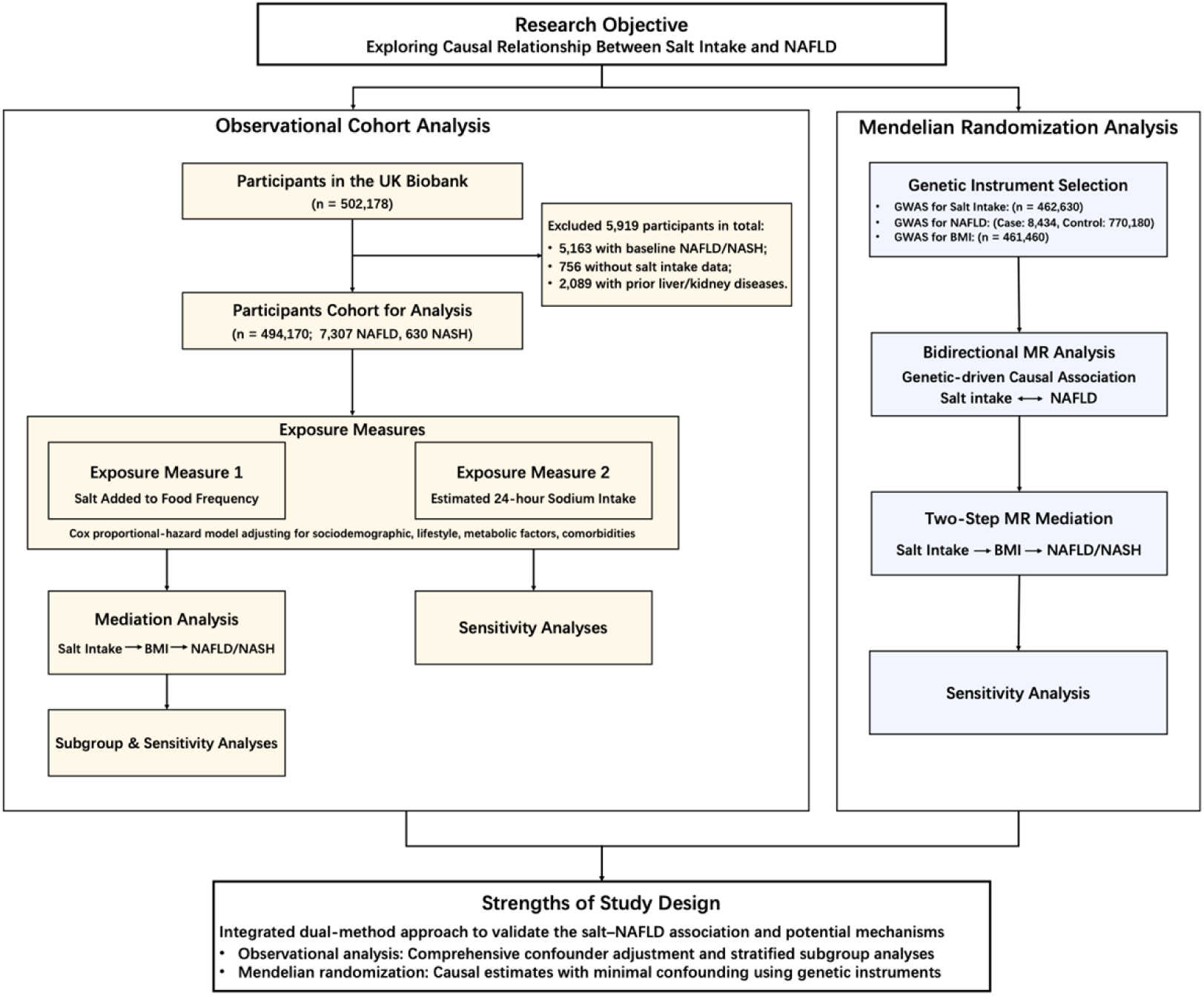
Illustration of study design.

## Methods

The study was approved by the UK Biobank, which has ethical approval from the North West Multi-center Research Ethics Committee in the United Kingdom. This analysis was conducted under UK Biobank application number 129053. All methods were reported in accordance with the Strengthening the Reporting of Observational Studies in Epidemiology–Nutritional Epidemiology (STROBE-nut) guidelines (S1 STROBE-nut Checklist), as well as the standard STROBE guidelines adapted for Mendelian randomization studies (S1 STROBE Checklist).

### Study population

The UK Biobank is a large, population-based prospective cohort study that enrolled more than half a million participants aged 37–73 years between 2006 and 2010 across Britain. For the current analysis, data from 494,170 participants were available. We excluded 5,163 individuals with a baseline diagnosis of NAFLD or NASH, 756 participants without salt intake data, and 2,089 individuals who had been diagnosed with any liver or kidney disease prior to baseline recruitment (Fig 1).

### Exposure assessment

Salt intake was assessed at baseline (2006–2010) using a touch-screen questionnaire [9]. Participants were asked, “Do you add salt to your foods? (Do not include salt used in cooking),” with four response options: (1) never/rarely, (2) sometimes, (3) usually, and (4) always. Individuals who selected "prefer not to answer" were excluded from the final analysis. Participants were also asked, “Have you made any major changes to your diet in the last 5 years?” with response options: (1) no; (2) yes, because of illness; and (3) yes, because of other reasons. This variable was used in sensitivity analyses to account for potential dietary modifications that could affect salt intake behavior.

In addition to self-reported behavior, we used estimated 24-hour urinary sodium excretion (mg/day) as a proxy for sodium intake, using the INTERSALT formula based on spot urine samples [10]. The sex-specific equations were:

- Sodium*_Men_* = 23 × (25.46 + 0.46×Na*_u_*− 2.75×Cr*_u_* − 0.13×K*_u_* + 4.10×BMI + 0.26×Age)
- Sodium*_Women_* = 23 × (5.07 + 0.34×Na*_u_*− 2.16×Cr*_u_* − 0.09×K*_u_* + 2.39×BMI + 2.35×Age − 0.03×Age^2^)

Where:

Na*_u_* = Urinary sodium (mmol/L)
Cr*_u_* = Urinary creatinine (mmol/L)
K*_u_* = Urinary potassium (mmol/L)
BMI = Body-mass index (kg/m²)
Age = Age at recruitment (years)

To assess the robustness of spot urine-based sodium estimates, we reviewed literature on the correlation between spot and 24-hour urine collections (r=0.6-0.8 in Western populations [11]). Sensitivity analyses using a correction factor for spot urine estimates were considered but not feasible due to lack of 24-hour urine data in the UK Biobank.

Given that BMI is a component of the INTERSALT formula, we conducted a sensitivity analysis using a modified version of the formula excluding BMI to minimize potential circularity in Cox models. Results remained directionally consistent with expected attenuation in effect sizes (S1 Table in S1 Appendix).

### Outcome identification

Incident NAFLD/NASH cases during follow-up were identified using the UK Biobank’s linked health records, including hospital admissions, primary care data, and death registries. According to the Expert Panel Consensus Statement [12], NAFLD (including NASH) was defined by the International Classification of Diseases Tenth Revisions (ICD-10) code K75.8 and K76.0. We further defined NASH by K75.8 as secondary outcomes. Follow-up time was calculated from the date of baseline assessment to the date of diagnosis, death, or censoring (January 1, 2024), whichever came first.

### Covariates assessment

Multiple potential confounders were adjusted for in the models, including sociodemographic characteristics, lifestyle factors, baseline comorbidities, and relevant biomarkers. Sociodemographic variables included age (continuous, in years), sex (male or female), ethnicity (White or non-White), and the Townsend Deprivation Index (TDI), a continuous composite measure of socioeconomic status derived from unemployment, non-car ownership, non-home ownership, and household overcrowding (with lower values indicating higher socioeconomic status).

Lifestyle factors included body mass index (BMI, continuous), smoking status (never, previous, or current), alcohol consumption (never, previous, or current), and healthy diet score (ranging from 0 to 5). The diet score was derived from intake thresholds of five dietary components (one score for each): vegetables (≥4 tablespoons/day), fruit (≥3 pieces/day), fish (≥2 times/week), unprocessed red meat (≤2 times/week), and processed meat (<2 times/week) [13, 14]. Physical activity level was categorized using the International Physical Activity Questionnaire (IPAQ, low, moderate, or high). IPAQ was further adjusted in a separate model given its 20.1% missing percentage. Sociodemographic, lifestyle, and dietary data were self-reported by participants.

Baseline comorbidities included type 2 diabetes and hypertension (yes/no), identified using ICD-10 codes (E11 and I10, respectively). Biomarkers included plasma triglycerides, alanine transaminase (ALT), glycated hemoglobin (HbA1c), and spot urinary potassium. Plasma biochemical markers such as triglycerides were measured using the Beckman Coulter AU5800 platform, while urinary potassium was analyzed via ion-selective electrode (ISE) technology on the AU5400 analyzer.

### GWAS and genetic instrument selection for Mendelian randomization

Genetic instruments for salt added to food and BMI were obtained from genome-wide association studies (GWAS) conducted within the UK Biobank study accessed via MRC IEU OpenGWAS platform [15]. These GWAS included 462,630 and 461,460 individuals of European ancestry, respectively, and were adjusted for age, sex, recruitment center, genotyping batch, and the first 10 principal components of genetic ancestry. Summary statistics for NAFLD were derived from a genome-wide meta-analysis of four electronic health record-based cohorts, including 8,434 cases and 770,180 controls of European descent [16].

#### Genetic instrument selection for salt-adding frequency

First, we selected single-nucleotide polymorphisms (SNPs) that were genome-wide significant (p<5*10^-8^) in the GWAS of salt-adding frequency. To ensure instrument validity and independence, we conducted linkage disequilibrium (LD) clumping using the clump_data function in the TwoSampleMR R package, applying a window of 10 Mb and *r*² < 0.001 [15]. Initially, 106 SNPs were identified as candidate instruments for salt-adding frequency. During harmonization with the NAFLD GWAS, 11 SNPs were unavailable and 7 SNPs replaced where possible with proxy SNPs (*r*² ≥ 0.8 within a 1 Mb window) using the European 1000 Genomes LD reference via the LDlink tool (https://ldlink.nci.nih.gov). Another 4 SNPs were excluded due to palindromic alleles with intermediate allele frequencies, yielding 98 final SNPs (MAF > 0.01) for the primary MR analysis (Data of Salt-NAFLD in S2 Appendix). The mean of F-statistics for the 98 SNPs were 51.0. Of these, 15 SNPs were identified as being associated with BMI through the PhenoScanner tool [17], and were excluded in sensitivity analyses to minimize the risk of horizontal pleiotropy.

#### Genetic instrument selection for NAFLD

For the reverse MR analysis, we similarly applied LD clumping using the clump_data function, with a 10 Mb window and an *r*² threshold of < 0.001. Four SNPs were identified as genome-wide significant and independent instruments for NAFLD and were available in the salt-adding frequency GWAS. None of these SNPs were palindromic with intermediate allele frequencies, avoiding strand ambiguity (Data of NAFLD-Salt in S2 Appendix). The mean F-statistic for the 4 SNPs used in the forward MR analysis was 90.87, indicating strong instrument strength. Among them, two were identified via the PhenoScanner tool as being associated with BMI and were excluded in a sensitivity analysis.

#### Genetic instrument selection for the multivariable MR analysis

Out of 1,767 SNPs that were strongly associated (p < 5×10⁻⁸) with either salt-adding frequency or BMI, 467 independent SNPs were retained after linkage disequilibrium (LD) clumping using a 10 Mb window and an *r*² threshold of < 0.001. During harmonization with the NAFLD GWAS, 76 SNPs were unavailable and 21 were replaced with proxy variants (*r*² ≥ 0.8 within a 1 Mb window), identified using the European 1000 Genomes LD reference panel via the LDlink tool. An additional 15 SNPs were excluded due to palindromic alleles with intermediate allele frequencies, resulting in 397 SNPs included in the final MVMR analysis (Data of MVMR in S2 Appendix). The conditional F-statistics were 55.25 for salt-adding frequency and 9.37 for BMI, indicating strong instrument strength for salt preference and moderate strength for BMI.

### Statistical analyses

#### Observational cohort statistical analyses

Baseline characteristics were summarized as mean (standard deviation, SD) for continuous variables and frequency (percentage) for categorical variables. Comparisons across salt intake frequency groups were conducted using chi-square tests for categorical variables and general linear models for continuous variables. Cox proportional hazards models were used to estimate hazard ratios (HRs) and 95% confidence intervals (CIs) for the association between salt intake frequency and incident NAFLD/NASH.

We also performed subgroup analyses and tested potential effect modification by following factors: sex, age (<60 or ≥60 years), BMI category (18.5–24.9, 25–29.9, ≥30 kg/m²), TDI quintiles (Q1, Q2–4, Q5), smoking and drinking status, baseline diabetes and hypertension, ALT (<40 or ≥40 U/L), triglyceride level (<1.7, 1.7–2.2, >2.2 mmol/L), HbA1c (<42 or ≥42 mmol/mol), and healthy diet score (low, moderate, high). To assess interactions between the frequency of adding salt to foods and these factors, multiplicative interaction terms were added to the Cox models and the Wald test was performed. Interactions were assessed by including product terms in the Cox models.

Given that weight loss remains the most evidence-based intervention for NAFLD [18], we evaluated whether BMI mediates the relationship between salt intake and NAFLD using a two-step mediation analysis based on VanderWeele’s framework [19]. As comparators, we also considered triglycerides and insulin resistance—proxied by the triglyceride-glucose (TyG) index, calculated as Ln (fasting triglycerides [mmol/L] x 88.57 x fasting glucose [mmol/L] x 18)/2 [20], due to their recognized roles in NAFLD [21–23].

We first assessed multicollinearity using variance inflation factors (VIF), with VIF >10 indicating collinearity. The mediation analysis proceeded under three conditions: (1) the exposure (salt-adding frequency) must be associated with the outcome (NAFLD); (2) the mediator (e.g., BMI) must be associated with the outcome; and (3) the inclusion of the mediator in the model should attenuate the salt–NAFLD association. Multivariable linear regression was used to estimate the effect of salt-adding frequency on BMI, and Cox proportional hazards models were used to assess associations between BMI and NAFLD risk, adjusting for the same covariates as in the primary model (excluding IPAQ due to missingness).

The indirect effect of salt intake on NAFLD via BMI was calculated using the product of coefficients method. Specifically, if the effect of salt-adding frequency on BMI is β₁ (SE = σ₁), and the effect of BMI on NAFLD is β₂ (SE = σ₂), the indirect effect θ₃ is estimated as: β_3_ = β_1_ × β_2_ with 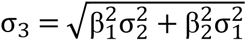. The proportion mediated was estimated as the ratio of the indirect effect to the total effect, with 95% CIs derived using 1,000 bootstrap replications [24].

Several sensitivity analyses were conducted to assess the robustness of the observational findings. First, to reduce potential reverse causality, we excluded NAFLD and NASH cases diagnosed within the first year of follow-up. Second, to account for potential changes in dietary habits influencing self-reported salt use, we excluded participants who reported modifying their diet in the past five years due to illness or other reasons. Third, we performed a sensitivity analysis restricted to participants with normal levels of ALT, BMI, and HbA1c, and without diabetes at baseline, as dose-response associations between salt intake and NAFLD were more pronounced among metabolically healthy individuals in subgroup analyses. Fourth, we conducted a sensitivity analysis for the BMI mediation analysis by excluding correlated covariates, including triglycerides and HbA1c.

#### Mendelian randomization statistical analyses

To assess the causal relationship between salt intake and the risk of NAFLD, we employed a MR framework. For valid causal inference, MR relies on three core assumptions: (1) Relevance—the genetic variants used as instruments must be robustly associated with the exposure of interest (e.g., salt-adding frequency or BMI); (2) Independence—these variants should not be associated with any unmeasured confounders of the exposure–outcome relationship; and (3) Exclusion restriction—the genetic instruments must influence the outcome (NAFLD) exclusively through the exposure, not via alternative pathways. While the relevance assumption can be statistically assessed, the independence and exclusion restriction assumptions cannot be directly tested and thus require careful methodological consideration and sensitivity analyses to support their plausibility.

Instrument strength was evaluated using the F-statistic, calculated as the square of SNP–exposure association (β²) divided by the square of its standard error (SE²) [25]. SNPs with F-statistics below the conventional threshold of 10 were excluded to avoid weak instrument bias. All SNPs were harmonized to align effect alleles for both salt-adding frequency and NAFLD. Inverse-variance weighting (IVW) with multiplicative random effects was used as main analysis, based on SNP-specific Wald estimates[26]. Sensitivity analyses using alternative MR methods included MR-Egger, weighted median, and MR-PRESSO to assess the robustness of results and detect potential horizontal pleiotropy [27, 28]. Where pleiotropy was evident, MR-Egger was prioritized for interpretation [29]. We further excluded BMI-associated SNPs to test the robustness of the estimates.

To validate the mediating role of BMI in the salt–NAFLD pathway, we performed a two-step MR mediation analysis. First, we estimated the effect of genetically predicted salt-adding frequency on BMI using IVW (β₁, σ_1_). Next, multivariable MR (MVMR) was used to estimate the direct effect of BMI on NAFLD, conditioning on salt intake (β₂, σ_2_), implemented using the MVMR R package [30]. Conditional F-statistics were calculated to assess instrument strength in the multivariable MR, and horizontal pleiotropy was tested using the modified Q-statistic [31]; MVMR-Egger was applied when horizontal pleiotropy was indicated. The indirect effect of genetically predicted salt-adding frequency on NAFLD mediated through BMI was calculated using the product of coefficients: β_3_ = β_1_ × β_2_[19], with its standard error (SE₃) estimated using the multivariate delta method: 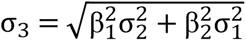. The mediation proportion was computed as the ratio of the indirect effect to the total effect, and 95% CIs were derived using the multivariate delta method [32].

All statistical analyses were performed using R version 4.2.4. All tests were two-sided, with a significance level set at *P* < 0.05.

## Results

### Participant characteristics

Among 494,170 participants (mean age 56.5 years; 45.6% male), 55.5% reported never or rarely adding salt to their food, 28.1% sometimes, 11.6% usually, and 4.8% always (Table 1). Compared to those with lower salt-use frequency, participants who reported adding salt more frequently were more likely to be of non-White ethnicity, have higher TDI scores and BMI, and were less likely to engage in healthy lifestyle behaviors (e.g., never smoking, high physical activity, or a high healthy diet score). Additionally, they had higher baseline prevalence of hypertension and diabetes, as well as elevated levels of serum HbA1c, ALT, and triglycerides. Correspondingly, both estimated 24-hour sodium intake and urinary potassium levels increased with salt-use frequency (Table 1). Self-reported salt-adding frequency correlated moderately with estimated 24-hour sodium intake (Spearman’s *r*=0.10, *P*<2.2*10^-16^), supporting its use as a proxy for sodium exposure, particularly reflecting long-term habitual intake (S1 Fig in S1 Appendix).

**Table 1.** Baseline characteristics according to the frequency of adding salt to foods.

| Characteristics | Total<br>(n=494,170) | Never/rarely<br>(n=274,146) | Sometimes<br>(n=138,631) | Usually<br>(n=57,480) | Always<br>(n=23,913) |
| --- | --- | --- | --- | --- | --- |
| Age, mean (SD), years | 56.53 (8.10) | 56.53 (8.08) | 56.43 (8.11) | 57.01 (8.03) | 55.95 (8.26) |
| Sex (male, %) | 225448 (45.6) | 120585 (44.0) | 63816 (46.0) | 29439 (51.2) | 11608 (48.5) |
| Ethnicity (White, %) | 468018 (95.0) | 262821 (96.2) | 130091 (94.1) | 53972 (94.2) | 21134 (88.8) |
| TDI, mean (SD) | -1.31 (3.08) | -1.50 (2.98) | -1.22 (3.11) | -1.09 (3.17) | -0.19 (3.50) |
| BMI, mean (SD), kg/m<sup>2</sup> | 27.42 (4.79) | 27.17 (4.72) | 27.63 (4.82) | 27.82 (4.81) | 28.04 (5.11) |
| Drinking |  |  |  |  |  |
| Never | 21825 (4.4) | 12065 (4.4) | 5657 (4.1) | 2358 (4.1) | 1745 (7.3) |
| Previous | 17435 (3.5) | 9974 (3.6) | 4354 (3.1) | 1914 (3.3) | 1193 (5.0) |
| Current | 454346 (92.0) | 251868 (92.0) | 128439 (92.8) | 53121 (92.6) | 20918 (87.7) |
| Smoking |  |  |  |  |  |
| Never | 269808 (54.8) | 162063 (59.3) | 72359 (52.4) | 25824 (45.1) | 9562 (40.2) |
| Previous | 170610 (34.7) | 89273 (32.7) | 50063 (36.3) | 22680 (39.6) | 8594 (36.2) |
| Current | 51879 (10.5) | 21892 (8.0) | 15638 (11.3) | 8735 (15.3) | 5614 (23.6) |
| Healthy diet score, mean (SD) | 2.87 (1.17) | 2.99 (1.15) | 2.82 (1.16) | 2.65 (1.17) | 2.42 (1.17) |
| IPAQ activity group |  |  |  |  |  |
| Low | 71633 (18.4) | 38444 (17.5) | 20206 (18.8) | 9066 (20.3) | 3917 (22.7) |
| Moderate | 158348 (40.7) | 90484 (41.2) | 43710 (40.6) | 17810 (39.8) | 6344 (36.7) |
| High | 159288 (40.9) | 90748 (41.3) | 43660 (40.6) | 17875 (39.9) | 7005 (40.6) |
| Baseline hypertension | 38031 (7.7) | 21493 (7.8) | 10357 (7.5) | 4235 (7.4) | 1946 (8.1) |
| Baseline diabetes | 6712 (1.4) | 3521 (1.3) | 1992 (1.4) | 823 (1.4) | 376 (1.6) |
| Baseline CVD | 78893 (16.0) | 43584 (15.9) | 21838 (15.8) | 9213 (16.0) | 4258 (17.8) |
| HbA1c, mean (SD), mmol/mol | 36.11 (6.73) | 35.94 (6.47) | 36.24 (6.91) | 36.35 (6.94) | 36.75 (7.94) |
| ALT, mean (SD), U/L | 23.51 (14.07) | 23.16 (13.63) | 23.78 (14.39) | 24.23 (14.47) | 24.17 (15.85) |
| TC, mean (SD), mmol/L | 1.75 (1.03) | 1.70 (0.99) | 1.77 (1.05) | 1.83 (1.09) | 1.86 (1.14) |
| UP, mean (SD), mmol/L | 63.10 (33.84) | 62.33 (33.71) | 63.68 (33.81) | 64.93 (34.20) | 64.09 (34.44) |
| Creatinine, mean (SD), mmol/L | 0.07 (0.01) | 0.07 (0.01) | 0.07 (0.01) | 0.07 (0.01) | 0.07 (0.01) |
| Sodium intake*, mean (SD), mg | 3500.48<br>(929.43) | 3419.10<br>(900.49) | 3551.18<br>(939.43) | 3673.65<br>(972.03) | 3724.88<br>(977.95) |

\*: Sodium intake was estimated by INTERSALT formula.
TDI, Townsend deprivation index; BMI, Body mass index; IPAQ, International Physical Activity Questionnaire; CVD, Cardiovascular disease; HbA1c, Glycated hemoglobin; ALT, Alanine transaminase; TC, Total triglycerides; UP, Urine potassium;

### Association between frequency of adding salt to foods and risk of NAFLD/NASH

During a mean follow-up of 14.3 years (7,077,709 person-years), 7,307 incident NAFLD cases (of these, 630 NASH cases) were recorded. After adjustment for age, sex, ethnicity, TDI, BMI, smoking and alcohol status, physical activity (IPAQ), healthy diet score, baseline hypertension and diabetes, triglycerides, HbA1c, and urinary potassium (model and covariate details: S2 Fig and S2 Table in S1 Appendix), a dose-response relationship was observed between salt-use frequency and NAFLD risk. Adjusted HRs (aHRs) across salt-use categories were 1.00 (reference), 1.02 (95% CI: 0.98–1.10), 1.20 (95% CI: 1.09–1.31), and 1.31 (95% CI: 1.16–1.48) across the groups of never/rarely, sometimes, usually, and always, respectively (*P* trend < 0.001, Table 2). For NASH, 630 incident cases were identified over 7,111,176 person-years. Participants who always added salt to food had a significantly higher risk of NASH compared to those who never or rarely added salt (aHR = 1.42; 95% CI: 1.01–1.99; S3 Table in S1 Appendix).

**Table 2.** The association between the frequency of adding salt to foods and hazard of NAFLD.

|  | Never/rarely | Sometimes | Usually | Always | *P*-trend |
| --- | --- | --- | --- | --- | --- |
| Case/total | 3,603/274,146 | 2,121/138,631 | 1,021/57,480 | 562/23,913 |  |
| Unadjusted | 1.00 (reference) | 1.17 (1.11-1.24) | 1.37 (1.28-1.47) | 1.84 (1.69-2.02) | <0.001 |
| Sex and age adjusted | 1.00 (reference) | 1.17 (1.11-1.24) | 1.36 (1.27-1.45) | 1.85 (1.69-2.02) | <0.001 |
| Multivariable adjusted<sup>a</sup> | 1.00 (reference) | 1.08 (1.02-1.15) | 1.23 (1.14-1.33) | 1.41 (1.27-1.56) | <0.001 |
| Multivariable adjusted + BMI<sup>b</sup> | 1.00 (reference) | 1.04 (0.98-1.11) | 1.18 (1.09-1.27) | 1.37 (1.24-1.52) | <0.001 |
| Multivariable adjusted + BMI + IPAQ<sup>c</sup> | 1.00 (reference) | 1.02 (0.98-1.10) | 1.20 (1.09-1.31) | 1.31 (1.16-1.48) | <0.001 |

<sup>a</sup>: Adjusted for age at recruitment, sex, ethnicity, smoking and drinking status, Townsend deprivation index, triglycerides, ALT, HbA1c, baseline diabetes and hypertension, potassium in urine and healthy diet score, 423,021 participants.
<sup>b</sup>: Further adjusted for BMI, 421,690 participants.
<sup>c</sup>: Further adjusted for IPAQ (International Physical Activity Questionnaire) activity group, 336,935 participants.

### Association between estimated 24-hour sodium intake and NAFLD/NASH risk

Each 1-gram increase in estimated 24-hour sodium intake was associated with increased risks of NAFLD (aHR = 1.82; 95% CI: 1.73–1.92) and NASH (aHR = 2.17; 95% CI: 1.81–2.61) after adjusting for major covariates (S4 Table in S1 Appendix). However, these associations were not statistically significant after further adjusting for BMI (aHR_NAFLD_: 1.07, 95% CI: 0.99-1.16; aHR_NASH_: 1.09, 95% CI: 0.83-1.46), suggesting potential mediation through adiposity. Sensitivity analysis by excluding BMI term in the INTERSALT formula showed consistent detrimental effects of higher sodium intake on NAFLD across all models (aHR_BMI-adjusted_: 1.07, 95% CI: 0.99-1.16; Table S1 in S1 Appendix).

### Mendelian randomization estimates of salt-adding frequency and NAFLD

MR analysis showed a positive association between genetically predicted frequency of adding salt to foods and NAFLD risk (Odds ratio, OR_IVW_ = 1.54; 95% CI: 1.16–2.05, Fig 2). This finding was consistent across sensitivity analyses using MR median and MR-PRESSO methods (Table 3). MR-Egger regression showed no evidence of horizontal pleiotropy, supporting the validity of the instrumental variables. Notably, the association remained robust after excluding SNPs associated with BMI or obesity (OR = 1.56; 95% CI: 1.18–2.06). Furthermore, bidirectional MR analysis did not support a reverse causal relationship from NAFLD to salt intake (OR = 1.01; 95% CI: 0.97–1.05). This null association remained consistent in sensitivity analyses after excluding the BMI-associated SNP (OR = 1.00; 95% CI: 0.99–1.02).

**Fig 2.**
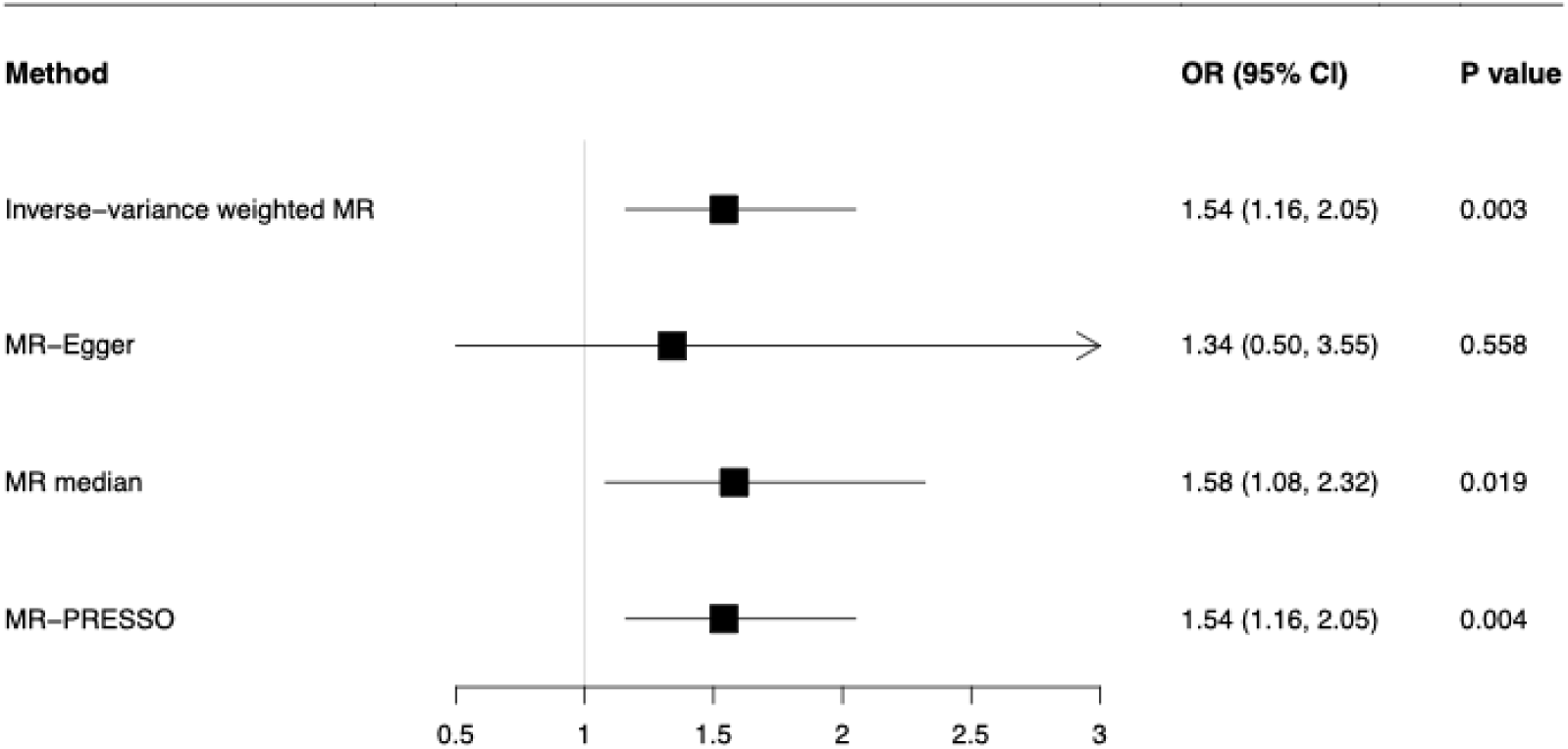
Mendelian randomization estimates of salt intake frequency and NAFLD risk. Forest plot showing MR estimates using inverse-variance weighted (IVW) and other sensitivity analyses for the genetic association of self-reported salt added to food with NAFLD risk.

**Table 3.** Mendelian randomization of adding salt to food and MAFLD.

| Method | Odds ratio | 95% CI | P value | P-intercept |
| --- | --- | --- | --- | --- |
| Inverse-variance weighted MR | 1.54 | 1.16, 2.05 | 0.003 |  |
| MR-Egger | 1.34 | 0.50, 3.55 | 0.558 | 0.765 |
| MR median | 1.58 | 1.08, 2.32 | 0.019 |  |
| MR-PRESSO | 1.54 | 1.16, 2.05 | 0.004 |  |

### Mediation of the salt-NAFLD association by BMI in the observational setting

In the observational setting, a higher frequency of adding salt to foods was significantly associated with higher BMI (β = 0.20; 95% CI: 0.18–0.22). In turn, BMI was positively associated with NAFLD/NASH risk in multivariable Cox models (aHR_NAFLD_ = 1.09; 95% CI: 1.08–1.10; aHR_NASH_ = 1.10; 95% CI: 1.07–1.14). Using a two-step bootstrap method for mediation analysis, we found that BMI partially mediated the association between salt-adding frequency and NAFLD/NASH, with mediation proportions of 16.4% (95% CI: 13.1–22.4%) and 18.1% (95% CI: 12.1–49.8%) for NAFLD and NASH, respectively (Fig 3). In contrast, we did not find significant mediation effects of triglycerides (1%, 95%CI: 0–2%) or TyG (1%, 95%CI: 1–3%). The lack of mediation by triglycerides or TyG index may reflect limited variability in these biomarkers at baseline or their downstream role in NAFLD progression. Future studies should measure dynamic changes in these markers over time to clarify their mediating effects.

**Fig 3.**
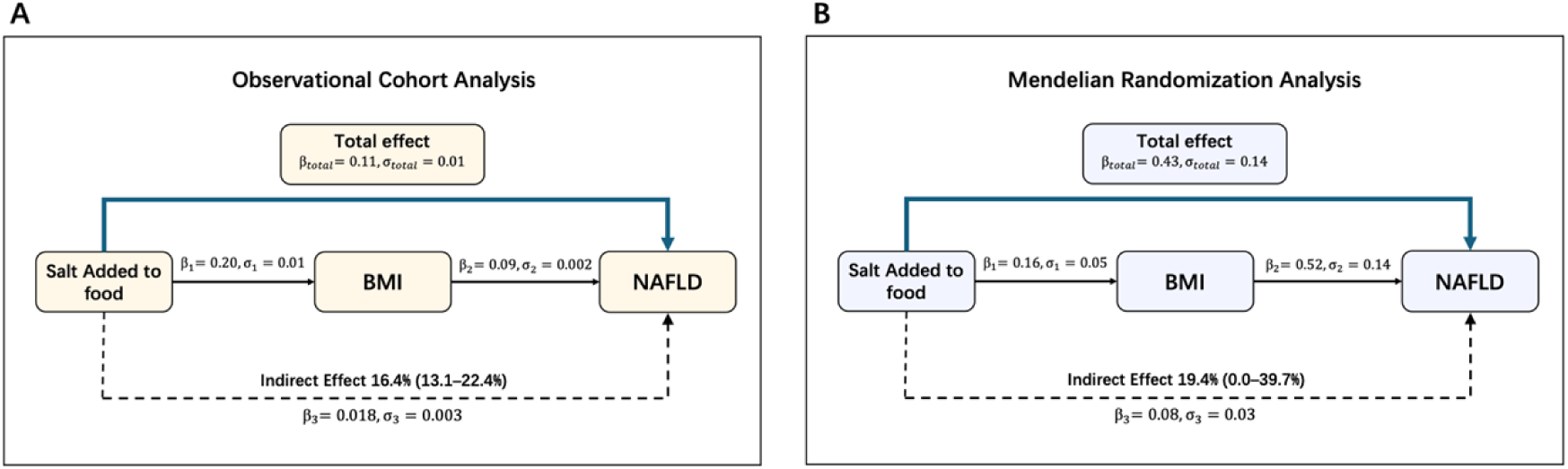
Mediation analysis of BMI on the association of salt intake frequency and NAFLD risk. Mediation analysis was applied on both observational (A) and Mendelian randomization (B) settings for the association of self-reported salt addition to food with NAFLD risk. The mediation proportions of BMI were similar for both approaches.

### Mediation of the salt-NAFLD association by BMI in MR analyses

In the MR mediation framework, genetic predicted salt-adding frequency was also positively associated with BMI (β_IVW_ = 0.16; 95% CI: 0.06–0.26; Figure 3). Accounting for potential horizontal pleiotropy (*P*_Q-statistic_<0.001), we used multivariable MR-Egger estimates, which confirmed a significant association between genetically predicted BMI and NAFLD risk (OR = 1.69; 95% CI: 1.29–2.21). The indirect effect of salt-adding frequency on NAFLD mediated through BMI yielded an OR of 1.09 (95% CI: 1.02–1.16). Two-step mediation suggested that BMI accounted for 19.4% (95% CI: 0.0–39.7%) of the total effect of salt-adding frequency on NAFLD (Fig 3), suggesting a modest but consistent mediating role across both observational and genetic analyses.

### Subgroup analysis

We assessed effect modification across 12 subgroups (e.g., age, sex, BMI, ALT, HbA1c) for the associations between salt-adding frequency and NAFLD risk (Table 4; Fig 4 and S3 Fig in S1 Appendix). The association was stronger among participants with BMI <25 kg/m², normal ALT (<40 IU/L), normal HbA1c (<42 mmol/mol), and no baseline diabetes (Fig 4). After applying false discovery rate (FDR) correction to account for multiple testing, BMI and ALT remained significant effect modifiers, confirming robust interactions.

**Fig 4.**
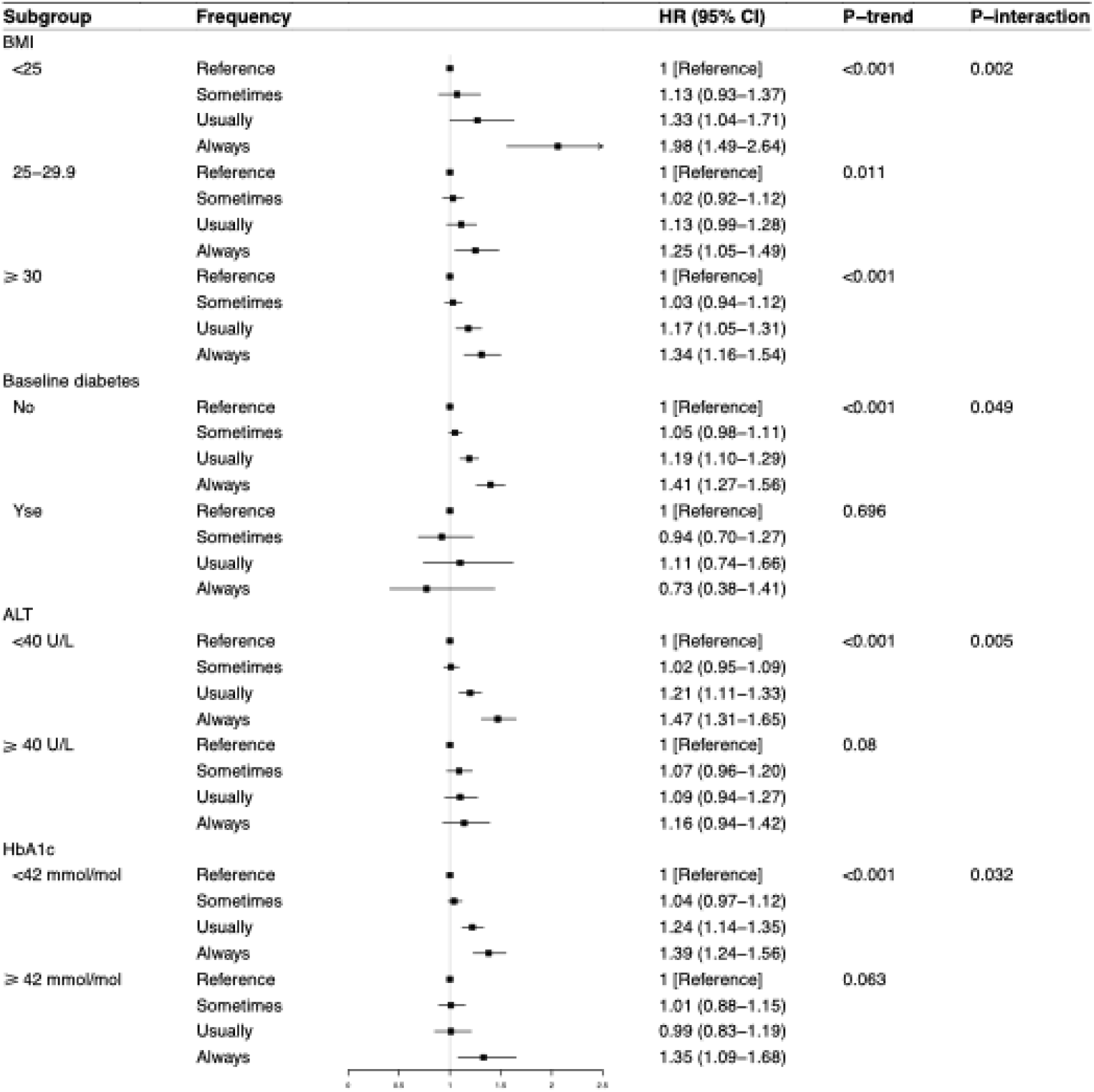
Subgroup analyses of salt-use frequency and NAFLD risk. Forest plot of stratified hazard ratios (HR) for the association between salt-use frequency and incident NAFLD, restricted to subgroups demonstrating significant effect modification. The effect modification by BMI and ALT remained significant after multiple comparisons.

**Table 4.** Subgroup analyses for the association of frequency of adding salt to food and risk of NAFLD.

| Subgroup | Never/<br>rarely | Sometimes | Usually | Always | *P*-trend | *P*-Interaction |
| --- | --- | --- | --- | --- | --- | --- |
| Age, y |  |  |  |  |  |  |
| <60 | 1 [Ref] | 1.07 (0.99-1.16) | 1.28 (1.16-1.42) | 1.44 (1.26-1.64) | <0.001 | 0.189 |
| ≥60 | 1 [Ref] | 1.00 (0.92-1.10) | 1.07 (0.95-1.20) | 1.29 (1.10-1.51) | 0.009 |  |
| Sex |  |  |  |  |  |  |
| Female | 1 [Ref] | 1.07 (0.98-1.16) | 1.15 (1.02-1.28) | 1.34 (1.16-1.54) | <0.001 | 0.653 |
| Male | 1 [Ref] | 1.01 (0.93-1.11) | 1.21 (1.09-1.35) | 1.39 (1.20-1.60) | <0.001 |  |
| BMI, kg/m² |  |  |  |  |  |  |
| <25 | 1 [Ref] | 1.07 (0.89-1.30) | 1.27 (1.00-1.63) | 2.06 (1.56-2.71) | <0.001 | 0.002 |
| 25-29.9 | 1 [Ref] | 1.03 (0.93-1.13) | 1.11 (0.97-1.26) | 1.25 (1.05-1.48) | 0.011 |  |
| ≥30 | 1 [Ref] | 1.03 (0.95-1.12) | 1.18 (1.06-1.31) | 1.31 (1.14-1.50) | <0.001 |  |
| Townsend deprivation index score, quantile |  |  |  |  |  |  |
| 1 (Low) | 1 [Ref] | 1.11 (0.95-1.29) | 1.09 (0.89-1.35) | 1.40 (1.03-1.90) | 0.041 | 0.134 |
| 2-4 (Intermediate) | 1 [Ref] | 1.06 (0.98-1.15) | 1.20 (1.08-1.33) | 1.51 (1.31-1.73) | <0.001 |  |
| 5 (High) | 1 [Ref] | 0.96 (0.86-1.08) | 1.17 (1.02-1.35) | 1.19 (1.01-1.41) | 0.011 |  |
| Smoking status |  |  |  |  |  |  |
| Never | 1 [Ref] | 0.97 (0.88-1.06) | 1.09 (0.96-1.23) | 1.44 (1.22-1.70) | 0.001 | 0.342 |
| Previous | 1 [Ref] | 1.09 (1.00-1.20) | 1.27 (1.13-1.43) | 1.30 (1.11-1.54) | <0.001 |  |
| Current | 1 [Ref] | 1.10 (0.94-1.29) | 1.15 (0.95-1.40) | 1.40 (1.14-1.73) | 0.002 |  |
| Drinking status |  |  |  |  |  |  |
| Never | 1 [Ref] | 0.94 (0.73-1.21) | 0.78 (0.53-1.14) | 1.19 (0.83-1.71) | 0.975 | 0.098 |
| Previous | 1 [Ref] | 0.83 (0.65-1.07) | 1.03 (0.74-1.42) | 1.26 (0.88-1.81) | 0.452 |  |
| Current | 1 [Ref] | 1.06 (1.00-1.13) | 1.21 (1.12-1.32) | 1.38 (1.24-1.54) | <0.001 |  |
| Baseline diabetes |  |  |  |  |  |  |
| No | 1 [Ref] | 1.05 (0.99-1.12) | 1.19 (1.10-1.28) | 1.40 (1.26-1.54) | <0.001 | 0.049 |
| Yes | 1 [Ref] | 0.92 (0.69-1.23) | 1.10 (0.74-1.62) | 0.77 (0.41-1.44) | 0.696 |  |
| Baseline hypertension |  |  |  |  |  |  |
| No | 1 [Ref] | 1.04 (0.97-1.11) | 1.18 (1.08-1.28) | 1.37 (1.22-1.53) | <0.001 | 0.950 |
| Yes | 1 [Ref] | 1.03 (0.89-1.18) | 1.18 (0.98-1.42) | 1.41 (1.12-1.78) | 0.004 |  |
| ALT level, U/L |  |  |  |  |  |  |
| <40 | 1 [Ref] | 1.01 (0.94-1.09) | 1.20 (1.09-1.31) | 1.47 (1.31-1.65) | <0.001 | 0.005 |
| ≥40 | 1 [Ref] | 1.09 (0.97-1.22) | 1.10 (0.95-1.27) | 1.14 (0.93-1.39) | 0.080 |  |
| Triglycerides, mmol/L |  |  |  |  |  |  |
| Normal | 1 [Ref] | 1.00 (0.91-1.10) | 1.21 (1.07-1.37) | 1.49 (1.27-1.74) | <0.001 | 0.269 |
| Borderline high | 1 [Ref] | 0.99 (0.86-1.13) | 1.19 (1.00-1.41) | 1.24 (0.99-1.56) | 0.023 |  |
| High | 1 [Ref] | 1.10 (1.00-1.20) | 1.15 (1.02-1.30) | 1.35 (1.15-1.57) | <0.001 |  |
| HbA1c, mmol/mol |  |  |  |  |  |  |
| <42 | 1 [Ref] | 1.04 (0.98-1.12) | 1.22 (1.12-1.33) | 1.38 (1.23-1.55) | <0.001 | 0.032 |
| ≥42 | 1 [Ref] | 1.01 (0.89-1.15) | 1.01 (0.85-1.21) | 1.33 (1.08-1.65) | 0.063 |  |
| Healthy diet score |  |  |  |  |  |  |
| Low (0-2) | 1 [Ref] | 0.98 (0.89-1.07) | 1.13 (1.01-1.27) | 1.32 (1.15-1.51) | <0.001 | 0.275 |
| High (3-5) | 1 [Ref] | 1.09 (1.01-1.18) | 1.21 (1.09-1.34) | 1.42 (1.22-1.64) | <0.001 |  |

### Sensitivity analyses

Findings remained consistent after excluding events occurring in the first year of follow-up or participants who had altered their diet in the past five years due to illness or other reasons (S5 & S6 Table in S1 Appendix). The association also persisted when analyses were restricted to metabolically healthy participants (defined by BMI <25 kg/m², ALT <40 IU/L and HbA1c<42 mmol/mol, and absence of diabetes at baseline; S7 Table in S1 Appendix). In a sensitivity analysis of the BMI mediation pathway, excluding correlated covariates (triglycerides and HbA1c) yielded a slightly higher but comparable mediation proportion of 20.3% (95% CI: 15.5–26.9%), supporting the robustness of BMI’s mediating role in the salt-NAFLD association.

## Discussion

In this large prospective cohort study, we observed that both a higher frequency of adding salt to foods and elevated estimated 24-hour sodium intake were associated with an increased risk of NAFLD/NASH, independent of socioeconomic status, lifestyle behaviors, dietary quality, and baseline comorbidities. These findings were further supported by Mendelian randomization analyses, which suggested a potential causal relationship between salt intake frequency and NAFLD risk. Notably, the association was more pronounced in metabolically healthy individuals, particularly those with normal BMI or ALT levels. Mediation analysis revealed that BMI partially mediated the salt-NAFLD relationship, suggesting adiposity as a mediator in the pathway linking excess salt consumption to fatty liver development.

Our results align with prior studies linking high sodium intake to metabolic syndrome, hepatic steatosis, and obesity-related complications [33–35]. We utilized two complementary measures—self-reported salt-adding frequency, reflecting long-term taste preference, and estimated 24-hour sodium intake, capturing hidden sodium from sources like processed foods. These findings are consistent with a Chinese cohort reporting increased NAFLD risk with high salt intake (≥10 g/day) [36] and a Korean cross-sectional study associating 24-hour urinary sodium with NAFLD and advanced fibrosis [37]. To minimize potential residual confounding, MR analysis further strengthened causal inference, showing that genetically predicted salt preference increased NAFLD risk, unaffected by BMI-related SNPs or reverse causation.

Several biological mechanisms may explain the observed associations. High salt intake has been shown to activate the aldose reductase–fructokinase pathway in the liver and hypothalamus, stimulating endogenous fructose production [38]. This process may induce leptin resistance and disrupt appetite regulation—a state in which satiety signaling is impaired—thereby promoting overeating and hepatic fat accumulation (Fig 5). These mechanisms align with our mediation analysis and previous evidence suggesting that obesity may act as a downstream mediator in the salt–NAFLD pathway. Supporting this, a bi-directional MR study indicated that salt intake causally increases BMI, whereas BMI does not influence salt-adding behavior, reinforcing its role as a mediator [39]. Moreover, excessive salt consumption has been associated with oxidative stress, insulin resistance, and dysregulated lipid metabolism—pathways known to contribute to hepatic fat accumulation and progression to steatosis and fibrosis [40, 41]. However, we did not observe a significant proportion of mediation by either insulin resistance indicator or triglyceride.

**Fig 5.**
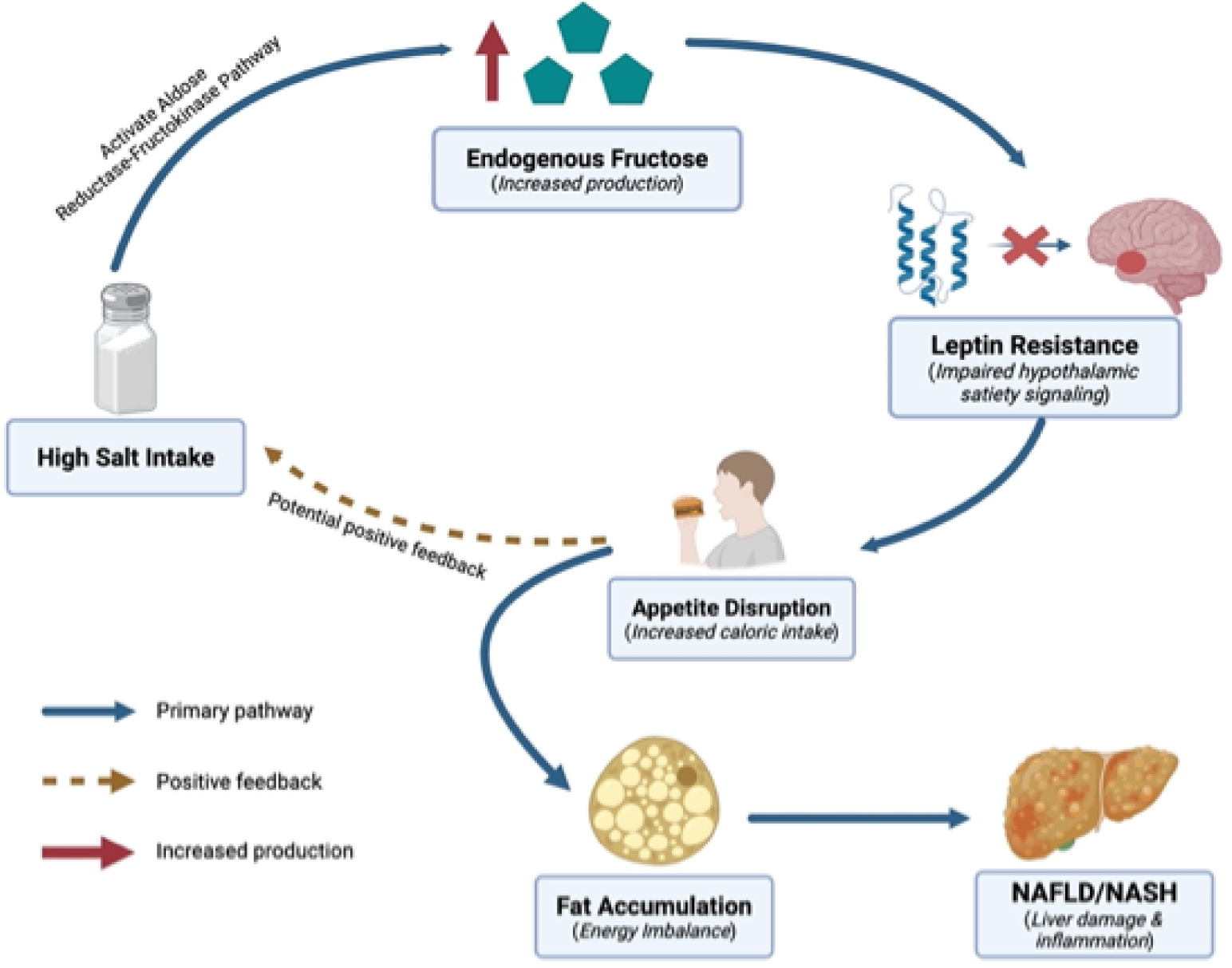
Proposed mechanisms linking salt intake to NAFLD/NASH. High salt intake may stimulate the aldose reductase–fructokinase pathway in both the liver and hypothalamus, leading to increased endogenous fructose production. This metabolic shift can induce leptin resistance, impairing satiety signaling and disrupting appetite regulation. The resulting increase in caloric intake may create a potential positive feedback loop, further elevating salt consumption through greater overall food intake. This energy imbalance promotes hepatic fat accumulation, ultimately driving the onset and progression of NAFLD and NASH.

Subgroup analyses revealed that the salt–NAFLD association was attenuated in individuals with higher BMI—a pattern also observed in previous studies examining salt intake in relation to chronic kidney disease and premature mortality [5, 42]. This attenuation may reflect a masking effect, whereby excess adiposity obscures the hepatic impact of salt. In contrast, the association was more pronounced and exhibited a clear dose-response pattern among individuals with normal ALT, HbA1c, and no baseline diabetes, suggesting that those without overt metabolic dysfunction may be particularly susceptible to the hepatotoxic effects of high sodium exposure.

To our knowledge, this is the first large-scale prospective study to comprehensively investigate the association between both behavioral (salt-adding frequency) and biochemical (estimated sodium intake) indicators of salt exposure and the incidence of NAFLD/NASH. By leveraging the extensive phenotypic, biochemical, and genetic data from the UK Biobank, our study design integrates the strengths of both observational epidemiology and genetic causal inference. The prospective cohort enabled rigorous adjustment for confounding variables and detailed subgroup analyses, while the Mendelian randomization approach provided complementary, minimally confounded estimates from a genetic perspective. This dual-method triangulation enhances the credibility of our findings by demonstrating consistent associations across distinct methodological frameworks and supports a potential causal pathway linking high salt intake to fatty liver disease through adiposity-related mechanisms.

Nonetheless, several limitations should be acknowledged. First, salt-use frequency was self-reported and may be subject to misclassification; however, the consistency of findings with biomarker-based sodium estimates supports the validity of this measure. Second, NAFLD and NASH diagnoses were derived from clinical coding and may underestimate true incidence. Third, although we adjusted for a comprehensive set of covariates, residual confounding from unmeasured or unknown factors cannot be entirely excluded in observational analyses; however, our Mendelian randomization approach helps mitigate this concern by providing less confounded estimates. Fourth, Spot urine-based sodium estimates may introduce measurement error, with studies suggesting 10-20% variability compared to 24-hour collections [11, 43]. This could attenuate associations, suggesting our findings may be conservative. Future studies should validate findings using 24-hour urine samples. Lastly, The UK Biobank’s predominantly White (95%) and middle-aged (37-73 years) cohort limits generalizability to other ethnic groups and younger populations. High salt intake is prevalent in Asian diets (e.g., via soy sauce, pickled foods), and NAFLD is rising in younger individuals [44]. Replication in diverse cohorts is needed.

Our findings support reducing salt intake to WHO’s recommended 5g/day (2g sodium), particularly in metabolically healthy individuals with normal BMI or ALT, who showed stronger salt-NAFLD associations. Dietary counseling targeting these groups could prevent early-stage NAFLD. Current NAFLD guidelines emphasize weight loss and exercise [45]. Public health policies, such as mandatory sodium labeling, reformulation of processed foods, and campaigns promoting low-sodium diets, could extend beyond cardiovascular benefits to reduce NAFLD risk. These strategies should prioritize populations with high salt consumption (e.g., processed food consumers). Our findings suggest adding salt reduction as a complementary strategy, particularly for early prevention in metabolically healthy individuals.

Our study provides converging evidence from both observational and genetic analyses that higher salt intake is associated with an increased risk of NAFLD and NASH. These findings expand the health implications of salt consumption beyond cardiovascular disease, highlighting a potential role in liver health. Public health strategies aimed at reducing salt intake may offer broader metabolic benefits, including the prevention of early-stage liver disease. Future trials and mechanistic studies are warranted to explore whether dietary salt reduction can modify NAFLD risk, particularly in metabolically healthy individuals. Future studies should incorporate imaging-based (e.g., ultrasound, MRI-PDFF) or biopsy-confirmed NAFLD/NASH diagnoses to improve diagnostic accuracy and capture subclinical cases, particularly for NASH, where underdiagnosis is likely. Mechanistic studies should measure biomarkers of the aldose reductase-fructokinase pathway (e.g., urinary fructose), leptin resistance (e.g., plasma leptin), and oxidative stress (e.g., malondialdehyde) to elucidate non-BMI pathways linking salt intake to NAFLD.

## Data Availability

Researchers registered with UK Biobank can apply for access to the UK Biobank Resource via portal (https://www.ukbiobank.ac.uk/). The summary data for all tables and figures are available in the appendix.

https://gwas.mrcieu.ac.uk/datasets/

## Acknowledgements

None.

## Supporting information captions

### Fundings

This study was funded by the National Key Research and Development Program of China (2023YFC2306902, 2023YFC2306900; to YYK and HY), the Project of the High-level Public Health Professional Talents of the Beijing Municipal Health Commission under Grant (XUEKEGUGAN-01-018; to YYK), and the Capital Medical University Cultivation Project (Natural Sciences Category; PYZ24073; to SL). The funders had no role in study design, data collection and analysis, decision to publish, or preparation of the manuscript.

### Authors’ contributions

Shun Li was responsible for conducting the study, performing data analysis, and drafting the manuscript. Hao Wang contributed to data collection and analysis. Xiaoqian Xu provided analytical support. Weiming Li was instrumental in developing and refining the figures. Hong You and Jidong Jia revised the manuscript. Youwen He contributed to data interpretation and conceptualization of the study. Yuanyuan Kong supervised the research and took overall responsibility for the study. All authors approved of the final version of the manuscript.

### Data reporting

The data that support the findings of this study are available in the UK Biobank at https://www.ukbiobank.ac.uk/ upon application.

### Related manuscripts

None.

